# Environmental contamination of the SARS-CoV-2 in healthcare premises: An urgent call for protection for healthcare workers

**DOI:** 10.1101/2020.03.11.20034546

**Authors:** Guangming Ye, Hualiang Lin, Liangjun Chen, Shichan Wang, Zhikun Zeng, Wei Wang, Shiyu Zhang, Terri Rebmann, Yirong Li, Zhenyu Pan, Zhonghua Yang, Ying Wang, Fubing Wang, Zhengmin (Min) Qian, Xinghuan Wang

**Affiliations:** Department of Laboratory Medicine, Zhongnan Hospital of Wuhan University, Wuhan, China; Department of Epidemiology, School of Public Health, Sun Yat-sen University, Guangzhou, China; Department of Epidemiology and Biostatistics, College for Public Health& Social Justice, Saint Louis University, USA; Department of Medical Administration, Zhongnan Hospital of Wuhan University, Wuhan, China; Department of Urology, Zhongnan Hospital of Wuhan University, Wuhan, China; Department of Nosocomial Infection, Zhongnan Hospital of Wuhan University, Wuhan, China; Center for Evidence-Based and Translational Medicine, Zhongnan Hospital of Wuhan University, Wuhan, China; Leishenshan Hospital, Wuhan, China

## Abstract

**Importance:** A large number of healthcare workers (HCWs) were infected by SARS-CoV-2 during the ongoing outbreak of COVID-19 in Wuhan, China. Hospitals are significant epicenters for the human-to-human transmission of the SARS-CoV-2 for HCWs, patients, and visitors. No data has been reported on the details of hospital environmental contamination status in the epicenter of Wuhan.

**Objective:** To investigate the extent to which SARS-CoV-2 contaminates healthcare settings, including to identify function zones of the hospital with the highest contamination levels and to identify the most contaminated objects, and personal protection equipment (PPE) in Wuhan, China.

**Design:** A field investigation was conducted to collect the surface swabs in various environments in the hospital and a laboratory experiment was conducted to examine the presence of the SARS-CoV-2 RNA.

**Setting:** Six hundred twenty-six surface samples were collected within the Zhongnan Medical Center in Wuhan, China in the mist of the COVID-19 outbreak between February 7 - February 27, 2020.

**Participants:** Dacron swabs were aseptically collected from the surfaces of 13 hospital function zones, five major objects, and three major personal protection equipment (PPE). The SARS-CoV-2 RNAs were detected by reverse transcription-PCR (RT-PCR).

**Main Outcomes and Measures:** SARS-CoV-2 RNAs

**Results:** The most contaminated zones were the intensive care unit specialized for taking care of novel coronavirus pneumonia (NCP) (31.9%), Obstetric Isolation Ward specialized for pregnant women with NCP (28.1%), and Isolation Ward for NCP (19.6%). We classified the 13 zones into four contamination levels. The most contaminated objects are self-service printers (20.0%), desktop/keyboard (16.8%), and doorknob (16.0%). Both hand sanitizer dispensers (20.3%) and gloves (15.4%) were most contaminated PPE.

**Conclusions and Relevance:** Many surfaces were contaminated with SARS-CoV-2 across the hospital in various patient care areas, commonly used objects, medical equipment, and PPE. The 13 hospital function zones were classified into four contamination levels. These findings emphasize the urgent need to ensure adequate environmental cleaning, strengthen infection prevention training, and improve infection prevention precautions among HCWs during the outbreak of COVID-19. The findings may have important implications for modifying and developing urgently needed policy to better protect healthcare workers during this ongoing pandemic of SARS-CoV-2.

**Key Points:** *Question:* What was the hospital setting contamination status, the most contaminated objects and PPE of SARS-CoV-2 during the outbreak of COVID-19 in Wuhan, China?

*Findings:* The most contaminated zones were the intensive care unit for novel coronavirus pneumonia (NCP) (31.9%), Obstetric Isolation Ward specialized for pregnant women with NCP (28.1%), and Isolation Ward for NCP (19.6%). The most contaminated objects and PPE are self-service printers (20.0%), hand sanitizer dispensers (20.3%), and gloves (15.4%).

*Meaning:* The findings may have important implications for modifying and developing urgently needed policy to better protect healthcare workers during this ongoing pandemic of SARS-CoV-2.

## Introduction

An outbreak of COVID-19 began in Wuhan, China in early December 2019 and is ongoing. There have been 80,955 confirmed COVID-19 cases, and 3,162 deaths in China as of March 11, 2020.^1^ Early on during the COVID-19 outbreak, healthcare workers (HCW) were found to beat high risk of developing COVID-19, even when infection prevention measures were in place, including usage of personal protective equipment (PPE: eye protection/face shield, respiratory protection, isolation gowns, and gloves), hand hygiene, and patient placement in negative-pressure isolation rooms. Thus far, 1,688 HCWs have become infected with SARS-CoV-2 in China, including 1,080 HCW cases in Wuhan where the outbreak started.^2,3^ However, the World Health Organization (WHO) indicates that many of the newer cases of HCW infection have stemmed from household exposures.^4^

The COVID-19 outbreak caused a sudden, significant increase in hospital visits from infected and suspected individuals over the course of two months.^5,6^ The large patient surge overwhelmed hospitals, despite continuous efforts to expand hospital capacity. Hospital waiting times were extended, which increased the time before infected individuals were identified and placed into isolation.

It is believed that the primary transmission mode of COVID-19 is through large respiratory droplets and close contact, although there is limited data that indicates that it may also spread through indirect contact with contaminated environments and aerosols.^4^ Only one study has examined possible environmental contamination of SARS-CoV-2 in a hospital outside of the epicenter of Wuhan and it consisted of a small sample size.^7^ Characterizing hospital contamination of SARS-CoV-2 is critical because hospitals have experienced massive patient surges during the outbreak and environmental contamination may contribute to disease spread.^8^ Data regarding the hospital function zones with highest levels of contamination can inform hospital cleaning and disinfection protocols to reduce the risk of healthcare-associated disease. The purposes of this study were to: (1) determine the extent to which the hospital environment becomes contaminated during outbreaks of COVID-19, (2)identify the highest areas of contamination within hospitals, and (3) identify the most frequently contaminated objects, medical supplies, and used PPE in a typical hospital in Wuhan, China during the ongoing outbreak of COVID-19.

## Methods

### Study location

The study was conducted in Zhongnan Medical Center of Wuhan University, located in Wuhan, China. Wuhan is the largest city in Central China and the capital city of Hubei Province in China.^9^ It has approximately 12 million people and has a subtropical, humid monsoon climate. Historically, Wuhan has been the biggest hub in China for land, water, and air transportation, and it is also one of the most industralized cities in China.^10^ The Zhongnan Medical Center possesses a Grade-III rating, the highest level according to a 3-tier system in China that recognizes a hospital’s capacity and ability to provide healthcare, conduct research, and deliver education. It has over 3,300 beds and a medical team that includes over 500 senior physicians. It includes 46 clinical departments and multiple research and clinical laboratories.

### Sampling

Samples were collected between February 7 - February 27, 2020, while the outbreak was ongoing. Three sets of surface samples were collected using dacron swabs across major hospital function zones, hospital equipment/objects and medical supplies, and HCW’s used PPE. Swab samples were also collected from control areas (i.e., spaces that did not house COVID-19 patients, consisting of administrative areas and the parking lot). Dacron swabs were premoistened with cell preservation solution. Samples were shipped with ice packs and refrigerated upon arrival at the laboratory. Blank controls were also used in swab sampling. Medical equipment assessed consisted of finger clips of pulse oximetry, electrocardiogram monitors, oxygen cylinders, oxygen regulators, oxygen masks, CT scanning machine, centrifuge, biosafety cabinet, and ventilator. Objects in non-medical areas (i.e., public facilities) assessed consisted of elevator buttons, microwave ovens, faucets, handrails, and hair drier. HCW’s’ used PPE assessed included gloves, eye protection or face shield and hand sanitizer dispensers; samples were collected after the HCW performed their duties with a COVID-19 patient. Hospital areas were classified into contamination zones, based on the percentage of swabs that were positive in that area/object.

### Reverse Transcription-PCR (RT-PCR)

Reverse transcription-PCR (RT-PCR) were conducted, using procedures recommended by the Chinese Center for Disease Control and Prevention. Briefly, we used the SARS-CoV-2 nucleic acid detection kits (DAAN Gene Co., Ltd, China) to extract viral RNAs.^11^ Two different targets on the SARS-CoV-2 genome were used: the ORF1ab and N genes. The Ct value of the amplification curve was defined as positive if less than 40 and negative if greater than 40. Both positive controls and negative controls were routinely included in each test.

### Statistical analysis

The R software (version 3.5.1) was used for all analyses. First, descriptive statistics were conducted. Differences in the positive detection rates of SARS-CoV-2 RNA in the surface swabs between hospital areas, hospital objects and medical supplies, and HCW’s used PPE were assessed using Chi-Square tests and/or the Fisher’s exact test where appropriate based on cell size. Two-tailed tests were used, and *P*-values smaller than 0.05 were considered statistically significant.

## Results

In total, 626 hospital environmental surface swab samples were collected; 13.6% were found to be positive for SARS-CoV-2 (Table 1). There were significant differences in the percentage of positive samples found across different hospital areas (*P* < .01). The most contaminated zones were the intensive care unit (ICU) that specialized in caring for NCP patients (31.9%), the Obstetric Isolation Ward focusing their care on pregnant women with NCP (28.1%), and the Isolation Ward for NCP patients (19.6%); these were classified as Contamination Zone III (CZ III). CZ II included the Outpatient Lobby (16.7% positive samples), Emergency Department (12.5%), Office and Preparation Area of the Isolation Ward for NCP patients (12.2%), Obstetric Ward (12.1%), and Clinical Laboratories (11.5%). CZ I included the Fever Clinics (6.5% positive samples), CT Examination Room (5.6%), and General Ward (5.5%). Both the Administrative Area and Parking Lot were classified as CZ 0.

**Table 1.** Percentage of Positive Hospital Environmental Samples for SARS-CoV-2 RNAs.

| Hospital Function Zone | Total Number, No. | Positive Number, No. (%) | *P* value |
| --- | --- | --- | --- |
| Total | 626 | 85 (13.6) | *P* < .001 |
| Contamination Zone III (High Positive Detection Rate) >18% |  |  |  |
| ICU <sup>a</sup> | 69 | 22 (31.9) |  |
| Obstetric Isolation Ward for NCP <sup>b</sup> | 32 | 9 (28.1) |  |
| Isolation Ward <sup>c</sup> | 56 | 11(19.6) |  |
| Contamination Zone II (Middle Positive Detection Rate), 9% - 18% |  |  |  |
| Outpatient Lobby | 30 | 5 (16.7) |  |
| Emergency Department | 80 | 10 (12.5) |  |
| Office and Preparation Area of the Isolation Ward for NCP | 41 | 5 (12.2) |  |
| Obstetric Ward | 33 | 4 (12.1) |  |
| Clinical Laboratories | 96 | 11(11.5) |  |
| Contamination Zone I (Low Positive Detection Rate) <9% |  |  |  |
| Fever Clinic | 46 | 3 (6.5) |  |
| CT Examination Room | 36 | 2 (5.6) |  |
| General Ward | 55 | 3 (5.5) |  |
| Contamination Zone 0 (Positive Detection Rate) = 0 |  |  |  |
| Administration Area | 42 | 0 (0) |  |
| Parking Lot | 10 | 0 (0) |  |

<sup>a</sup>The intensive care unit for NCP only.
<sup>b</sup>The ward for the pregnant women who were diagnosed novel coronavirus pneumonia (NCP).
<sup>c</sup>The isolation ward specialized in caring for NCP patients.

Among all examined commonly used hospital objects and medical equipment, 13.9% were found to be positive for SARS-CoV-2 (Table 2). This included positive samples from self-service printers (20%, it is a machine commonly used in China by patients themselves to print out their examination or test reports in a hospital), desktops (16.8%), doorknobs (16.0%), and telephones (12.5%), medical equipment (12.5%), and public facilities (8%). Only 5.6% of samples collected from walls and floors were positive. The most contaminated objects were self-service printers (20%), desktops/keyboards (16.8%), and doorknobs (16%).

**Table 2.**
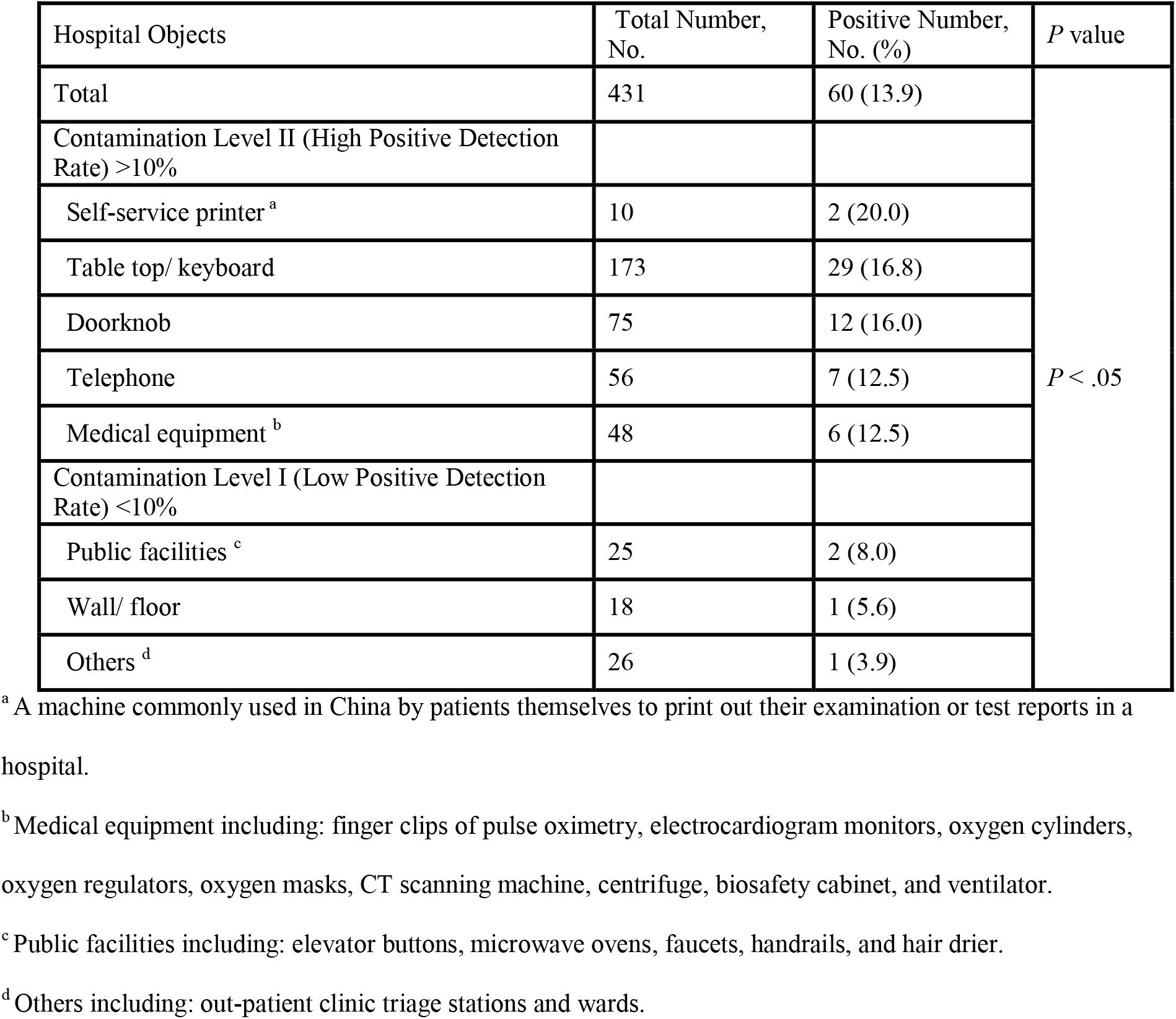
Percentage of Positive Hospital Object Samples for SARS-CoV-2 RNAs.

Of the samples collected from HCWs’ used PPE (hand sanitizer dispensers, gloves, and eye protector/face shield), 12.9% were positive for SARS-CoV-2 (Table 3). Significant differences were found in the percentage of positive samples across the PPE types (*P* < .01). The highest positive detection rate found was from the hand sanitizer dispensers 20.3%; 15.4% and 1.7% of gloves and eye protection or face shields tested positive, respectively (Table 3).

**Table 3.** Percentage of Positive Samples of SARS-CoV-2 RNAs for Health Care Workers’ Used Personal Protection Equipment.

| Personal Protection Equipment (PPE) | Total Number, No. | Positive Number, No. (%) | *P* value |
| --- | --- | --- | --- |
| Total | 195 | 25 (12.8) | *P* < .01 |
| Hand sanitizer Dispensers | 59 | 12 (20.3) |  |
| Gloves | 78 | 12 (15.4) |  |
| Eye Protection or Face Shield | 58 | 1 (1.7) |  |

## Discussion

This investigation showed that the hospital environment frequently becomes contaminated when providing care to COVID-19 patients. Contaminated areas/items included patient care areas housing COVID-19 patients, common hospital objects/items, such as self-service printers, desktops, doorknobs, and keyboards, medical equipment, and HCWs’ PPE, such as gloves, eye protection, and face shields. These findings suggest that the hospital environment could potentially be a source of virus spread, including among HCWs, patients, and visitors. It is noteworthy that the hospital surface samples were collected from February 7 to February 27, 2020, which was after human-to-human spread of SARS-CoV-2 was identified. It is plausible to assume that the hospital surface contamination would have been more severe before the sampling period, when environmental cleaning protocols were not as extensive and HCWs were not aware of the potential risk of indirect contact spread. HCWs may have unwittingly spread the virus throughout the hospital via contaminated hands, PPE, and equipment. Direct or indirect contact with these potentially highly contaminated surfaces may account for the first wave of patients into the hospital and the early cases of healthcare associated transmission among HCWs and visitors.

The WHO-China Joint Mission on Coronavirus Disease 2019 (COVID-19) Report stated that HCW occupational exposure to SARS-CoV-2 may not play a significant role in transmission, based on the limited cases investigated.^12^ However, the data in this study suggests that hospital environmental contamination with SARS-CoV-2 is extensive, which could be an important occupational risk for HCWs. Generally speaking, HCWs have better infection prevention knowledge and practice compared to general populations. Despite this, 2,055 infected HCWs have been confirmed from healthcare facilities across China by February 20, 2020. These exposures may have been related to multiple factors. Hospitals were providing care to COVID-19 patients for over a month before human-to-human spread was formally identified as one important transmission mode on January 20, 2020. During this time, widespread hospital contamination from SARS-CoV-2 could have led to occupational exposure for HCWs working in the hospital setting on a daily basis. Immediately after recognizing that SARS-CoV-2 was spreading through human-to-human, the hospital became overcrowded by a massive patient surge. This sudden, unusually high demand for medical care far exceeded the hospital’s capacity. Hospital waiting time extended to hours and families commonly had to seek care at multiple facilities before they could be seen. This increased the time before infected individuals were identified and placed into isolation, which could have contributed to healthcare-associated disease transmission. In addition, during the early stages of the outbreak, HCWs did not always follow standard infection prevention measures due to a lack of medical supplies, especially in areas not designated as infection-specific units. For example, critical infection prevention measures, such as wearing respiratory and eye protection, gowns, and gloves, were not instituted for HCWs in non-infection units. Furthermore, the study hospital experienced a sudden transition from a comprehensive medical center to a focused care center for COVID-I9 infected and confirmed patients in order to respond to the patient surge. This transition occurred in a matter of days. The majority of HCWs in the hospital had no direct experience with this type of outbreak, and a significant proportion HCWs in non-infection units did not receive infection prevention training during the outbreak. These sudden changes may have led to higher exposure to SARS-CoV-2 for HCWs as well as causing massive hospital surface contamination fromSARS-CoV-2.

In this study, the most contaminated hospital zones included the ICU that specialized in caring for COVID-I9 patients, the Obstetric Isolation Ward only taking care of pregnant women with COVID-I9 infection, and the Isolation Ward for COVID-I9 patients. It was surprising to observe the highest positive detection rate of SARS-CoV-2 in the isolation ward for pregnant women diagnosed with COVID-I9. In China, it is a custom for multiple family members and friends to visit pregnant women around the time of delivery and the average hospitalization for pregnant women giving birth usually lasts more than five days. Isolation of pregnant women and new mothers during hospitalization has been challenging and often impossible. Family members and friends infected with or carrying SARS-CoV-2 could have contaminated the obstetric isolation ward during their visits. This finding emphasizes the need for strict isolation practices that should be implemented rapidly. In addition, pregnant women and neonates are highly susceptible populations. They are more sensitive to the chemicals released by the internal infection control processes. It is, therefore, a standardize procedure to lessen the intensity and to reduce frequency of the internal infection control processes in the department, which may have led to the high positive detection rates discovered from our study. It was not surprising that the Fever Clinic had a low positive detection rate for SARS-CoV-2 (6.5%). Early on, the Fever Clinic was relocated to a semi-open area, which was well-ventilated. In addition, unlike the obstetric isolation ward, strict infection prevention measures were instituted in the Fever Clinic. These practices likely led to the low positive detection rate.

It is notable that both hand sanitizer dispensers and HCWs’ used gloves possessed the highest positive detection rate of SARS-CoV-2 compared to eye protection or face shields. Gloves would become contaminated by touching infected patients and/or contaminated surfaces in the hospital. Eye protection and face shields would likely become contaminated by infected patients’ respiratory droplets or infectious aerosols (if produced by COVID-19), or by the HCWs’ own hands during donning or doffing. This study’s data supports the idea that contaminated gloves or hands may be a possible exposure route for SARS-CoV-2. Patients and visitors usually do not wear gloves in hospital settings. Contaminated hands and surfaces could potentially lead to SARS-CoV-2 exposure. Hospitals should emphasize the critical role of hand hygiene as a way to prevent SARS-CoV-2 infection.

## Limitations

This study has several limitations. SARS-CoV-2 culture samples were not collected; thus, exact bioburden levels were unable to be determined. No air samples were collected during the investigation. This study focused on hospital surface contamination of SARS-CoV-2 RNAs as a surrogate of exposure to SARS-CoV-2. We were limited in our ability to characterize other exposure factors, such as other exposure routes, frequencies, and duration. A lack of resources also meant that we were unable to conduct a comprehensive exposure study for the HCWs working in the hospital in the midst of the ongoing outbreak.

## Conclusions

Many environmental surfaces were contaminated with COIVD-19 RNA across the hospital in various patient care areas, commonly used objects, medical equipment, and PPE. The contamination could be caused by viral shedding from infected patients and/or indirect contact by HCWs, patients, and visitors. These findings emphasize the need to ensure adequate environmental cleaning, strengthen infection prevention training, and improve infection prevention precautions among HCWs during the outbreak of COVID-19.

## Data Availability

The reported data are available from the corresponding authors on reasonable request. After publication of the findings, the data will be available for others upon the request. Our team will provide contact information including an email address for future communication once the data are ready to be shared with others. The detailed study plan will be needed for assessment of the reasonability to request for the data. The corresponding authors will make a decision based on the provided documents. Additional information may also be needed during the process.

## ARTICLE INFORMATION

### Author Affiliations

Center for Evidence-Based and Translational Medicine, Department of Urology, Zhongnan Hospital of Wuhan University, Leishenshan Hospital (Wang);Department of Laboratory Medicine, Zhongnan Hospital of Wuhan University (Ye, Chen, Wang, Zeng, Wang, Li, Wang); Department of Epidemiology and Biostatistics, College for Public Health & Social Justice, Saint Louis University (Qian and Rebmann); Department of Epidemiology, School of Public Health, Sun Yat-sen University (Lin and Zhang); Department of Medical Administration, Zhongnan Hospital of Wuhan University (Pan);Department of Nosocomial Infection, Zhongnan Hospital of Wuhan university (Wang); Department of Urology, Zhongnan Hospital of Wuhan University (Yang)

### Author Contributions

Ye, Wang had roles in the study design, clinical management, data collection, data analysis, data interpretation, literature search, and writing of the manuscript. Wang, Qian had roles in the study design, clinical management, data analysis, data interpretation, literature search, and writing of the manuscript. Lin, Rebmann had roles in the data analysis, data interpretation, literature search, and writing of the manuscript. Chen, Wang, Zeng, Wang had roles in data collection, data analyses, literature search, and intensive manuscript revision. Li, Pang, Yang, Wang had roles in the clinical management, data collection, data analysis. All authors reviewed and approved the final version of the manuscript.

## Acknowledgements

We wanted to express our highest appreciation to our staff who risked their lives collecting the large number of samples needed to conduct this study. We also thank the nurses and doctors in Zhongnan Medical Center for their support and assistance for collecting the samples.

